# Comparative analysis of policies and programs to support families and children during COVID-19

**DOI:** 10.1101/2021.03.04.21252943

**Authors:** Joanne Kearon, Mark Cachia, Sarah Carsley, Meta van den Heuvel, Jessica Hopkins

**Author notes:** **Corresponding Author** Jessica Hopkins. **Author’s Contribution:** JK, SC, MvdH and JH conceptualized the project and assisted in creation of the search strategy. JK, MAC, MvdH, and JH performed data extraction. All authors contribute to data analysis. JK drafted the manuscript. SC, MvdH and JH contributed in editing the manuscript. All authors approved the final manuscript.

## Abstract

**Background:** Policies and programs that promote positive social environments for young children and their families have the potential to improve early childhood development and long-term health. However, due to the community-wide public health measures implemented to reduce transmission of COVID-19, many families are experiencing health and socio-economic challenges and pre-existing supports and services may no longer be available. In this study, we compared the policies and programs countries have implemented to support maternal and child health during the first wave of COVID-19.

**Methods:** We compared the policies and programs implemented to support child health and well-being during the first wave of COVID-19 in Australia, Canada, the Netherlands, Singapore, the UK, and the USA. A grey literature review was performed to identify policies, announcements, and guidelines released from governmental and public health organizations within each country related to children, parents, families, early childhood development, adverse childhood experiences, child welfare, pre-school, or daycares. We also performed a manual search of government websites. Both provincial and federal government policies were included for Canada.

**Results:** The main policies identified were focused on prenatal care, well-baby visit and immunization schedules, financial supports, domestic violence and housing, childcare supports, child protective services, and food security. All of the included countries implemented some of these policies, but there was a large variation in the number, size, and barriers to access these supports. None of the countries implemented supports in all of the potential areas identified.

**Conclusion:** Political legacy and previous redistributive policies might have influenced the variation in policies and programs introduced by governments. As the COVID-19 pandemic continues, further opportunity for governments to implement supportive programs and policies for children and families exists.

## Background

Healthy growth and development, from the prenatal environment to the first 5 years of life, plays a major role in health and well-being across the life course (Maggi et al., 2010). Environmental factors during early childhood influences the probability of future obesity, heart disease, mental health, educational outcomes and involvement in the criminal justice system (Hertzman, 2010). In particular, adverse childhood experiences (ACEs) occurring in the first 18 years of life are associated with long-term stress-related changes in the nervous, endocrine and immune systems, leading to multiple poor health conditions such as type 2 diabetes and cognitive decline (Danese & McEwen, 2012).

Thus, policies that promote positive social environments for young children and their families have the potential to improve early childhood development, and subsequently health for entire populations. For example, across several countries from a range of political legacies, those that provided mothers with longer maternal leaves and higher maternal pays also had higher rates of breastfeeding initiation (van den Heuvel et al., 2013). Similarly, countries that publicly funded early childhood education centres had higher literacy rates at 15 years of age than those that were privately funded (van den Heuvel et al., 2013). Furthermore, programs that target specific populations or subgroups can decrease social inequity within countries (Navarro et al., 2006; Siddiqi et al., 2011).

There is growing concern about the impact of COVID-19 and the community-wide public health measures to reduce transmission of COVID-19 (eg. quarantine, daycare closures) on child health, parent health and family functioning (Thapa et al., 2020; Van Lancker & Parolin, 2020). COVID-19 has drastically changed how public health services, prenatal, neonatal and pediatric care are delivered. As such, many of the policies created to support early childhood development have been more difficult to access or implement during COVID-19. Several papers have considered how health and social services can be delivered safely during the pandemic without diminishing access, quality and effectiveness of care (Bogler & Bogler, 2020; Graham et al., 2020; Jago et al., 2020; Zhang et al., 2020). Publications to date have highlighted gaps that already existed in supporting families in some countries (Hynan, 2020; Thapa et al., 2020), and how the pandemic may widen inequities in prenatal and obstetric care (Onwuzurike et al., 2020). Daycares and schools often serve as a mechanism to provide food to low-income children at risk of food insecurity, and other health services (eg. vaccinations, mental health services). With school closure during COVID-19, alternative systems needed to be found (Dunn et al., 2020). Recommendations have been made on policies that can mitigate the impact of COVID-19 on children, such as reducing barriers to accessing social support, providing supports for women at risk of domestic violence, and increased support for education for children that lack technology resources (Douglas et al., 2020).

The objective of this paper is to compare the policies and supports that different countries have put into place to support child health during COVID-19. We will then explore the reasons for potential varying approaches, as well as discuss the potential impact of varying approaches on health and well-being.

## Methods

We aimed to answer the question, “How did governments support child health during the COVID-19 pandemic through policies and programs?” We compared the policies and programs created by governments in selected countries during their first wave of COVID-19.

### Country Selection

The following countries were included in this study: Canada (including the provinces of British Columbia, Alberta, Ontario and Quebec), United Kingdom (UK), Australia, Netherlands, United States of America (USA), and Singapore. Country selection was discussed and finalized amongst all authors. The focus was on the Canadian experience, with information being gathered about both federal and provincial policy interventions. We then selected Organization of Economic Cooperation and Development (OECD) comparator countries representing a variety of political legacies and COVID-19 epidemiologic experiences (Canadian Institute for Health Information, 2018). Within the USA, in addition to federal policies being explored, Michigan was also chosen for further comparison due to its similarities to Ontario, for which it has previously been used as a comparator to Ontario (R. P. Murphy et al., 2016).

### Baseline Country Characteristics and COVID-19 Epidemiology

Baseline information about each country was gathered, including population, Gross Domestic Product (GDP) per capita, percentage (%) of GDP spent on early childhood education (ECE), enrollment of children 3-5 years in ECE, immunization coverage, and Gini coefficient. Gini coefficient is a measure of income inequality within a group of people, on a range from 0 to 1, with higher numbers indicating greater inequality(De Maio, 2007). The main sources of data were the World Bank (2021) and OECD (2006, 2019, 2020), though in some cases, country specific reports were used due to lack of synthesized international data.

We also compared the countries’ experiences of the COVID-19 pandemic during their first wave by collecting data on date of first case, date of the first peak in cases, the incidence during the peak, their testing capacity during the first wave, and the Government Stringency Index (GSI) during their peak, which is a measure of the intensity of a country’s community-wide public health measures. This information was found through Our World in Data (Ritchie et al., 2021c), a project based at the University of Oxford.

### Search Strategy and Data Sources

The search strategy was designed to identify policies, announcements, and guidelines released from governmental and public health organizations within each country related to children, parents, families, early childhood development, adverse childhood experiences, child welfare, pre-school, or daycares. Specifically, we were interested in clinical guidelines, financial benefits or government funding of health or social programs, and childcare supports. Therefore, we focused on a search of the grey literature.

We designed and reviewed the search strategy with a Public Health Ontario health sciences librarian. We devised search strings based on our targeted policy topics and repeated the search on a range of pre-designed search engines that pulled information specifically from governmental and public health bodies, as well as on Google. For each search string on each search engine, the first 100 results for each search were reviewed. Based on the findings of this search, targeted iterative manual searches on Google were also completed. Following this, we conducted a manual search of each country’s governmental websites to ensure that no other policy decisions were missed by the original search strategy. A manual search of the Netherland’s policies was completed by an author fluent in the language and then translated to English. We also reviewed guidance documents from national and provincial/state clinical colleges or bodies in pediatrics, obstetrics, and family medicine. The search was performed from August to November 2020. The full search strategy is outlined in Appendix A.

We chose to focus on pre-school-aged children (0-5 years), and therefore any policies or programs specific to school-aged children were excluded (though policies that would apply to both pre-school- and school-aged children were included). As well, policies that were open to the general public, but not specifically targeting children, parents or families, were excluded. We included policies introduced by these governments from January to August 2020.

## Results

### Baseline Characteristics of Selected Countries

Table 1 provides an overview of the baseline characteristics of the included countries. In terms of population size, the range was from Singapore’s population of 5.9 million to USA’s 328.2 million (World Bank, 2021). While Singapore has the smallest population, it has the highest population density. The GDP per capita ranged from $42 330.10 (USD) in the UK to $65 297.50 (USD) in the USA, compared to $46 194.70 (USD) in Canada (World Bank, 2021). In this group of countries, the Gini coefficient ranged from 0.285 in the Netherlands to 0.458 in Singapore (Li, 2020; OECD, 2020a).

**Table 1.** Baseline characteristics of included countries

| | Population<br>(millions) | Population<br>Density (/km<sup>2</sup>) | GDP per capita<br>(\$USD) | % of GDP Spent<br>on ECE | Enrollment of<br>children 3-5 years<br>old in ECE (%) | Measles<br>immunization<br>coverage (%) | Gini<br>coefficient |
| --- | --- | --- | --- | --- | --- | --- | --- |
| Australia | 25.4 | 3 | 55 060.30 | 0.66 | 84 | 95 | 0.325 |
| Canada | 37.6 | 4 | 46 194.70 | 0.20 | 24 | 90 | 0.303 |
| Netherlands | 17.3 | 511 | 52 331.30 | 0.60 | 95 | 94 | 0.285 |
| Singapore | 5.9 | 7953 | 65 233.30 | 0.19 | 84 | 95 | 0.458 |
| UK | 66.8 | 275 | 42 330.10 | 0.65 | 100 | 91 | 0.366 |
| USA | 328.2 | 36 | 65 297.50 | 0.33 | 66.1 | 90 | 0.390 |

We also aimed to provide baseline information about the current status of child health and supports for children in the country. In some cases, there was heterogeneity in the way the data was reported between countries, and the year of the most recent data available. The amount spent on ECE, as a percentage of GDP, ranged from about 0.20% in Singapore in 2011 and Canada in 2006, to over 0.65% in the UK and Australia in 2015 (Early Childhood Development Agency, 2012; OECD, 2006, 2019). The percentage of children 3-5 years old enrolled in ECE ranged from 24% in Canada in 2006 to 100% in the UK in 2017 (OECD, 2006, 2019). Immunization coverage for all of the childhood vaccines in these countries was >90%. The largest differences were seen with measles vaccine coverage, which ranged from 90% in Canada and the USA, to 95% in Australia and the UK (Vanderslott et al., 2013).

### COVID-19 Epidemiologic Indicators During First Wave

An overview of the epidemiology of COVID-19 in the included countries can be seen in Table 2. All of the countries included identified their first case in late January or early February 2020, with the exception of the Netherlands, which identified their first case in late February. The date of the first wave peak ranged from late March to late July (Ritchie et al., 2021a). At peak, the number of cases per day ranged from 15.0 cases/million people in Australia to 203.5 cases/million people in the USA (Ritchie et al., 2021a). GSI scores indicate that public health measures implemented during each country’s peak were most relaxed in the USA, and the most stringent in Singapore (Ritchie et al., 2021c). Testing capacity and test positivity at peak was also assessed. Australia had the second highest testing capacity (2.62 per 1000), but the lowest test positivity (0.7%), while the USA had the highest testing capacity (2.89 per 1000), but mid-range test positivity (8.9%) (Ritchie et al., 2021b). On the other hand, the Netherlands, Singapore and the UK had lower testing capacities, and high test positivity rates (19.1-28.9%) (Ritchie et al., 2021b). Canada testing capacity was 0.7 per 1000 with a test positivity of 6.6% at the peak of the first wave (Ritchie et al., 2021b).

**Table 2.** Comparison of epidemiology of COVID-19 in included countries until August 31, 2020

|  | Date of 1<sup>st</sup><br>reported<br>case | Date of peak<br>new cases per<br>day | Peak new cases per<br>day/million | GSI at date of peak<br>new cases | Testing capacity per 1000<br>people at peak new cases | Test positivity<br>at peak (%) |
| --- | --- | --- | --- | --- | --- | --- |
| Australia | January 25 | March 30 | 15.0 | 79.17 | 2.62 | 0.7 |
| Canada | January 26 | May 4 | 47.7 | 72.69 | 0.7 | 6.6 |
| Netherlands | February 28 | April 15 | 65.4 | 79.63 | 0.33 | 21.5 |
| Singapore | January 24 | April 27 | 171.8 | 85.19 | 0.54 | 28.9 |
| UK | February 1 | April 24 | 71.4 | 79.63 | 0.37 | 19.1 |
| USA | January 21 | July 23 | 203.5 | 67.13 | 2.89 | 8.9 |

### Healthcare: Prenatal, Neonatal and Pediatric Care

Table 3 gives an overview of the overarching recommendations and guidance from clinical professional agencies and organizations from each country related to prenatal, neonatal and pediatric care.

**Table 3.** Comparison of policies regarding prenatal and pediatric care in selected countries during COVID-19

| Country | Jurisdiction | Prenatal Care | Well-baby visit schedule | Vaccines |
| --- | --- | --- | --- | --- |
| Australia | Federal | Continue routine antenatal care, though provider can arrange for extra scans if COVID-19 positive | Continue routine schedule with mix of virtual and in-person visits | Recommendation to continue vaccinations as scheduled |
| Canada | Federal | Modified schedule, with mix of virtual and in-person practice<br>Consider delaying routine appointments for pregnant patients being tested or COVID-19 positive | Modified schedule with mix of virtual and in-person visits | Recommendation to continue vaccinations as scheduled |
|  | Alberta | Prenatal classes suspended | Public health nurse or midwife to continue postpartum care as usual<br>Physician visits should continue, but may be virtual depending on location | Routine immunizations to continue, with exception of school program |
|  | British Columbia | Reduced antenatal visits, with a mix of virtual and in-person practice | At the discretion of physician | Routine immunization schedule to continue |
|  | Ontario | Modified prenatal visit schedule | Modified well-child schedule, with mix of in-person and virtual | Routine infant vaccination schedule<br>Consider delaying 4-6 year old immunizations |
|  | Quebec | Modified schedule, with mix of virtual and in-person practice | Routine schedule to be continued, though may be virtual | Routine infant vaccination schedule |
| Netherlands |  | Continue routine schedule | Combination of in person and virtual visits<br>In-person weight checks by appointment only if there are concerns | Routine schedule to continue |
| Singapore |  | Postpone non-critical appointments if on a Stay-Home Notice or quarantine<br>Otherwise, continue routine schedule |  | Continue routine schedule |
| UK |  | • Continue routine antenatal care, although can be modified, unless suspected or confirmed COVID-19 | • Continue routine 6-8 week infant examination | • Continue routine childhood vaccinations as scheduled |
| USA | Federal | • Continue to provide medically necessary prenatal care, referrals and consultations but can modify/reduce if risk outweighs benefit | • Continue with routine well-baby visits | • Recommendation to continue infant and toddler vaccinations as scheduled |
|  | Michigan | • Policy varied by health service provider<br>• General reduction in in-person visits | • At the discretion of physician | • Continue routine schedule |

Overall, there was general uniformity within the recommendations across countries, with only small variations. Each clinical body provided information on infection prevention and control (IPAC) policies, or links to related resources. As well, most clinical guidance focused on the clinical treatment of COVID-19 in patients (eg. pregnant women, infants). Of particular relevance was the guidance provided to health care providers on how to alter patient care schedules in order to reduce in-person contact. Most clinical organizations recommended continuing with current care schedules (ie. prenatal care, well-baby care). Canadian organizations recommended integrating virtual appointments when possible into the schedule, both for prenatal and pediatric care (Bogler & Bogler, 2020; Elwood et al., 2020). As well, all countries’ clinical bodies recommended some flexibility in the care schedule if the patient were to have COVID-19 or be suspected to have COVID-19 - in those situations, a delay of 2 weeks would be permissible. Only Singapore recommended prenatal appointments be delayed if there was a general lockdown (College of Obstetricians & Gynaecologists, Singapore, 2020). The USA recommended that the prenatal schedule be reduced if risk outweighs benefit (American College of Obstetricians and Gynecologists, 2020). Finally, there was agreement that the infant series of vaccination should continue as scheduled.

There were a few other notable variations. For example, most jurisdictions specified that only one support person could accompany a patient during labour, while the Netherlands and Quebec allowed for two (Government of Quebec, 2020a; Nederlandese Vereniging Voor Obstetrie & Gynaecologie, 2020). As well, there was variation in the degree that different clinical organizations or agencies recommended that individual healthcare providers perform their own risk assessment based on their location and patient.

### Maternal Supports

Table 4 provides an overview of governmental policies and programs to support mothers and/or parents.

**Table 4.**
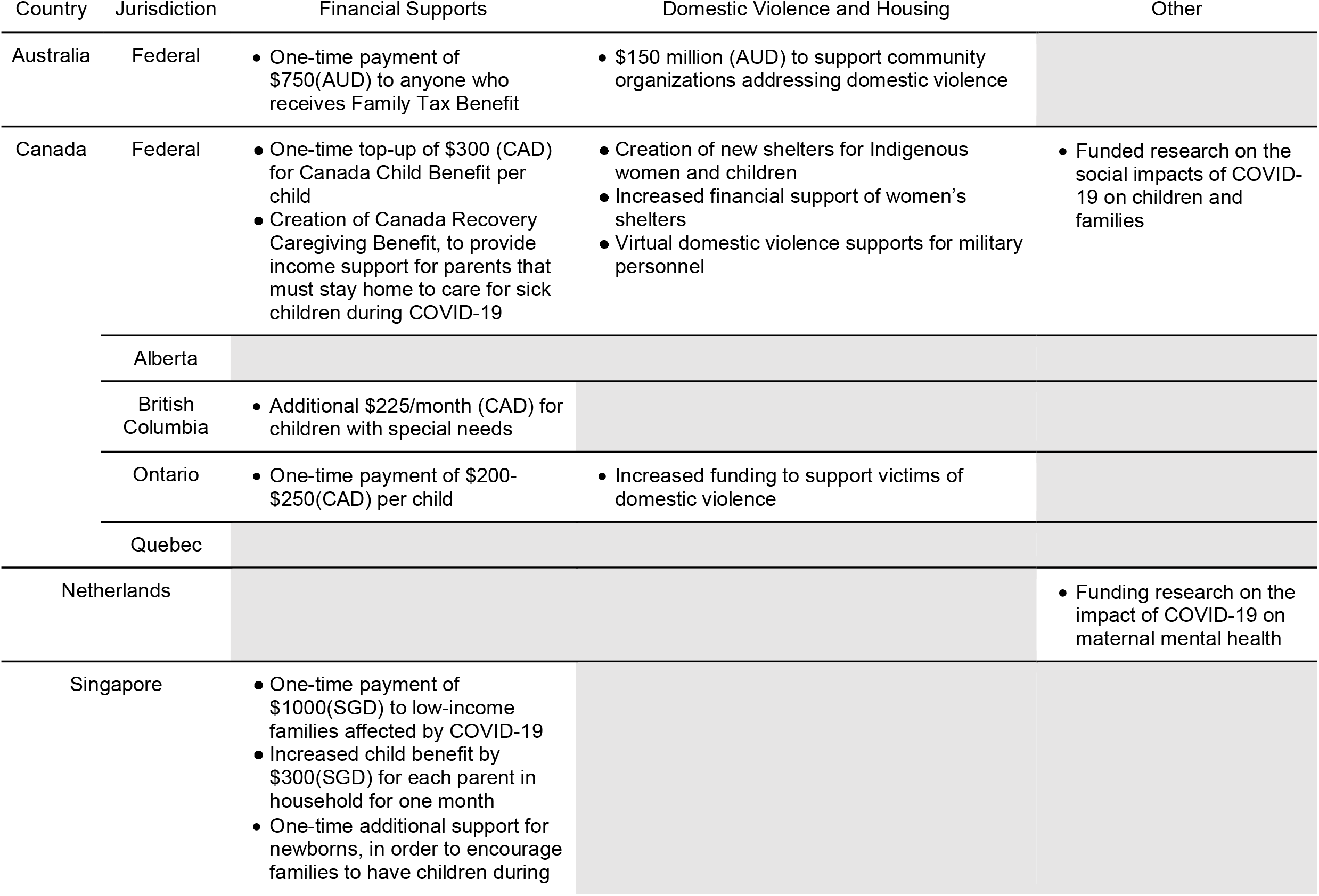
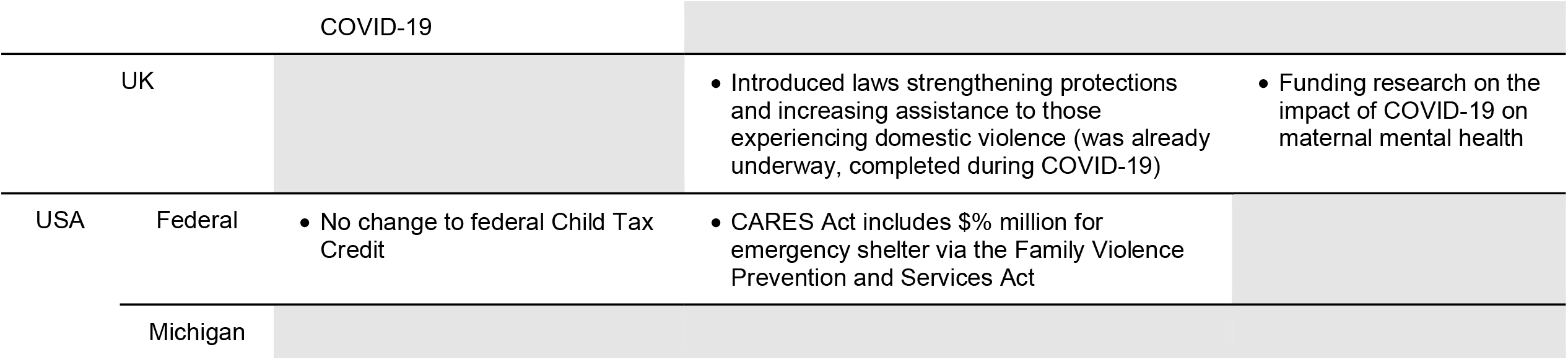
Comparison of additional maternal supports offered by governments in response to COVID-19 in selected countries

In terms of financial supports, Canada, Australia and Singapore created specific benefits for parents, in addition to pre-existing supports, as well as financial supports that were offered to the general population during COVID-19 (Budget 2020, Government of Singapore, 2020; Canada Revenue Agency, Government of Canada, 2020a; Department of Social Services, Australian Government, 2020). Within Canada, British Columbia provided an additional $225(CAN)/month benefit for children with special needs (Ministry of Children and Family Development, Government of British Columbia, 2020), and Ontario provided a one-time $200-$250(CAN) payment per child additional to the federal benefit (Ministry of Education, Government of Ontario, 2020). Notably, there was variation in how these benefits can be accessed, with Canada and Australia both requiring application to the benefits, while Singapore provided the benefit automatically to anyone who was already receiving other child benefits (Budget 2020, Government of Singapore, 2020; Canada Revenue Agency, Government of Canada, 2020a; Department of Social Services, Australian Government, 2020). Canada introduced an additional financial support in August 2020 for parents who are forced to take time off of work in order to care for a child that must isolate(Canada Revenue Agency, Government of Canada, 2020b). Singapore also provided an additional separate benefit for low-income families, and a benefit for newborns (Budget 2020, Government of Singapore, 2020). This latter program was explicitly created to encourage people to continue having children during the COVID-19 pandemic. The UK, Netherlands and USA did not make any changes to their current policies or programs related to financial support for families.

Canada and Australia both increased funding to community organizations addressing domestic violence and for women’s shelters (Canada, 2020; K. Murphy, 2020). Additionally, Canada’s federal government created additional shelters specifically for Indigenous women and children (Canada, 2020). During the COVID-19 pandemic, the UK passed a law that increased protections and supports for those experiencing domestic violence (Home Office, UK Government, 2020). However, that law was already being considered and drafted prior to the emergence of COVID-19.

Several countries, including Canada, UK and Netherlands, specifically funded research related to the impact of COVID-19 on women and children (Canadian Institutes of Health Research, Government of Canada, 2020; Dutch Research Council (NWO), 2020; Economic and Social Research Council, 2020).

All of the countries’ governmental websites provided general information related to maternal mental health and well-being. As well, all countries’ governmental websites provided links to community organizations, programs, or other supports that already exist to aid parents and young children.

### Childcare and Early Child Development

Table 5 provides an overview of each countries’ governments’ support for early childhood development including approaches to childcare, child protective services and food security.

**Table 5.**
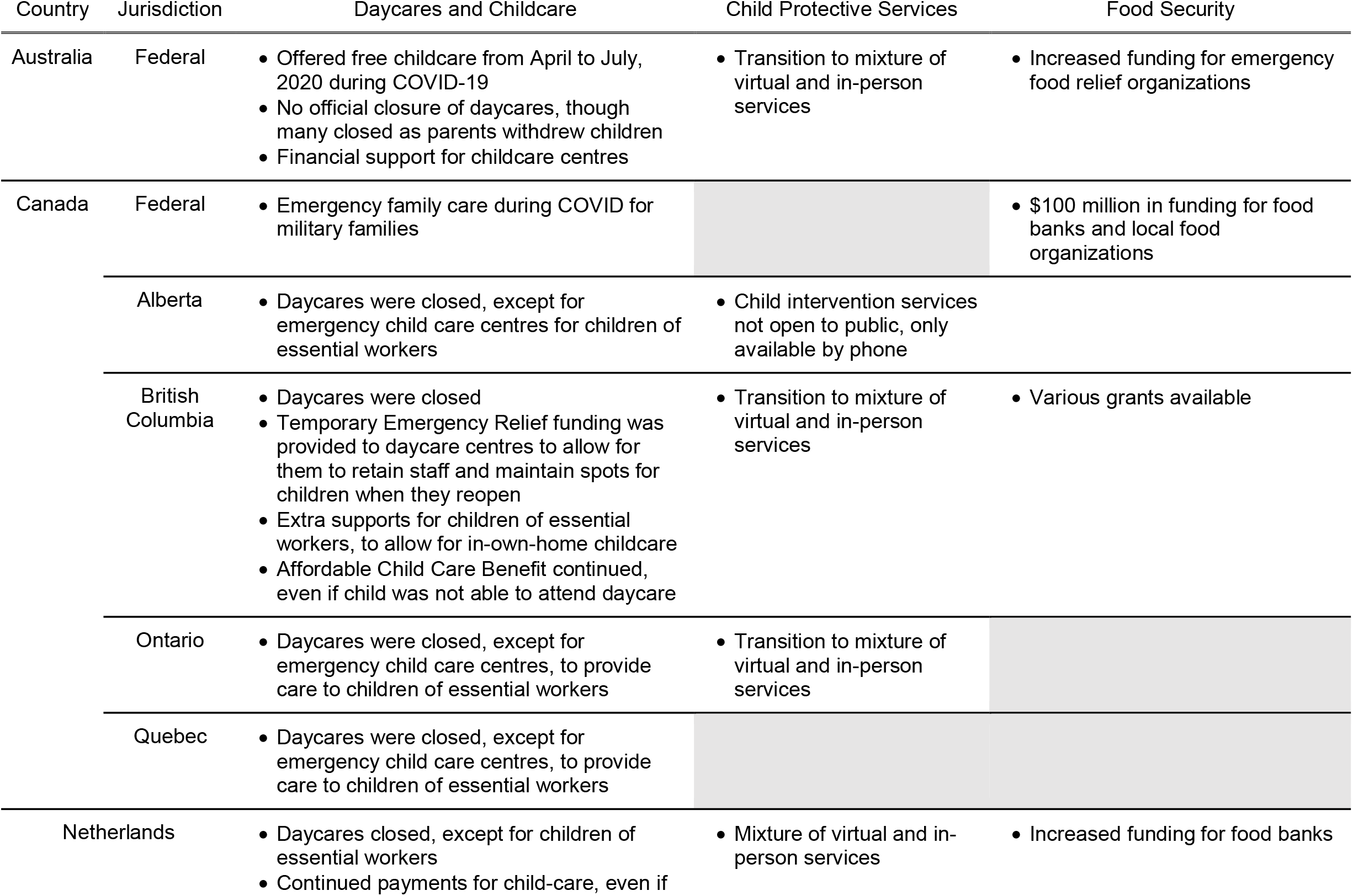
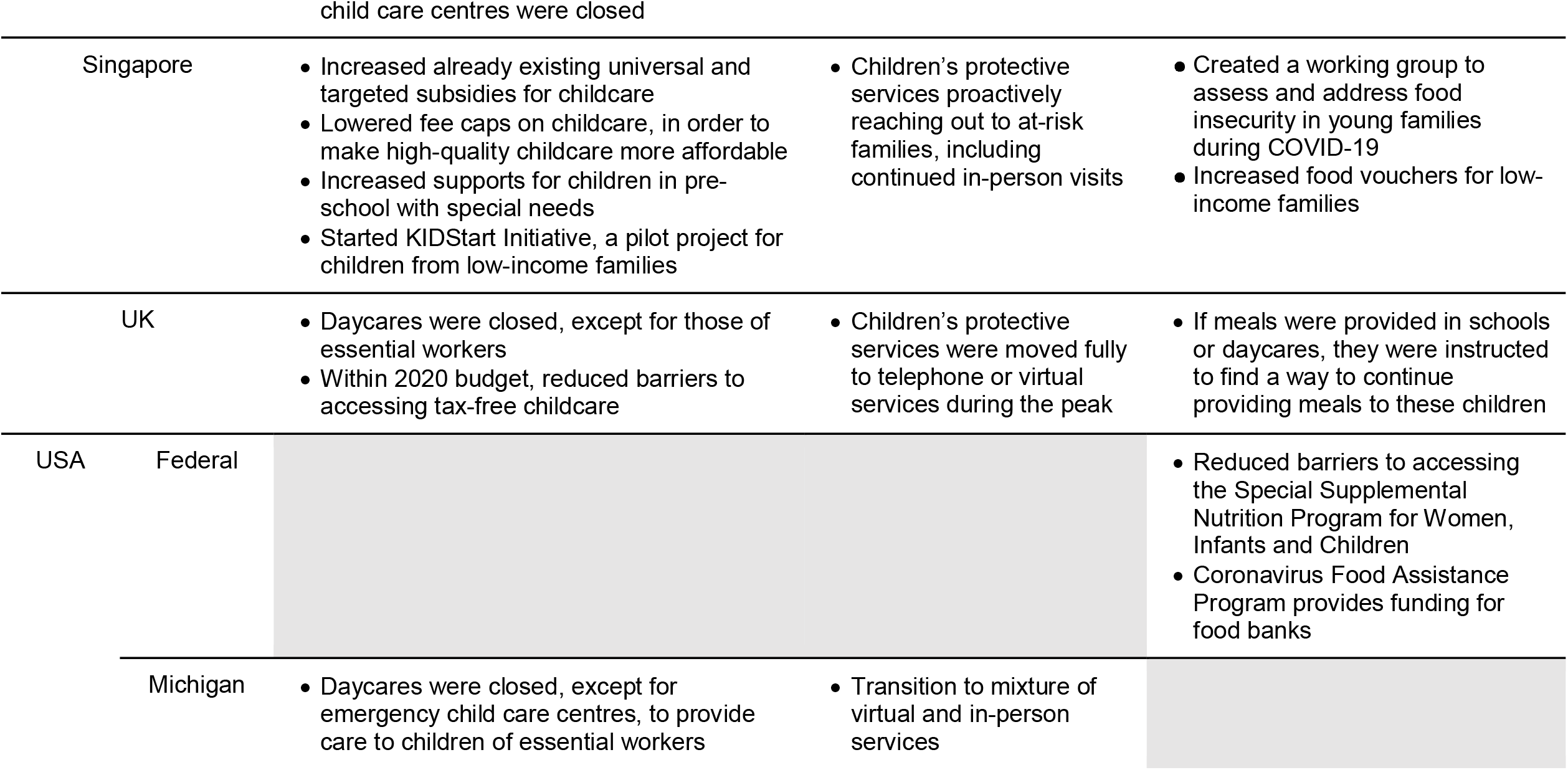
Comparison of additional supports for childcare and early childhood development by governments in response to COVID-19 in selected countries

| Country | Jurisdiction | Daycares and Childcare | Child Protective Services | Food Security |
| --- | --- | --- | --- | --- |
| Australia | Federal | • Offered free childcare from April to July, 2020 during COVID-19<br>• No official closure of daycares, though many closed as parents withdrew children<br>• Financial support for childcare centres | • Transition to mixture of virtual and in-person services | • Increased funding for emergency food relief organizations |
| Canada | Federal | • Emergency family care during COVID for military families | | • \$100 million in funding for food banks and local food organizations |
|  | Alberta | • Daycares were closed, except for emergency child care centres for children of essential workers | • Child intervention services not open to public, only available by phone |  |
|  | British Columbia | • Daycares were closed<br>• Temporary Emergency Relief funding was provided to daycare centres to allow for them to retain staff and maintain spots for children when they reopen<br>• Extra supports for children of essential workers, to allow for in-own-home childcare<br>• Affordable Child Care Benefit continued, even if child was not able to attend daycare | • Transition to mixture of virtual and in-person services | • Various grants available |
|  | Ontario | • Daycares were closed, except for emergency child care centres, to provide care to children of essential workers | • Transition to mixture of virtual and in-person services |  |
|  | Quebec | • Daycares were closed, except for emergency child care centres, to provide care to children of essential workers |  |  |
| Netherlands |  | • Daycares closed, except for children of essential workers<br>• Continued payments for child-care, even if | • Mixture of virtual and in-person services | • Increased funding for food banks |
| Singapore |  | • Increased already existing universal and targeted subsidies for childcare<br>• Lowered fee caps on childcare, in order to make high-quality childcare more affordable<br>• Increased supports for children in pre-school with special needs<br>• Started KIDStart Initiative, a pilot project for children from low-income families | • Children's protective services proactively reaching out to at-risk families, including continued in-person visits | • Created a working group to assess and address food insecurity in young families during COVID-19<br>• Increased food vouchers for low-income families |
| UK |  | • Daycares were closed, except for those of essential workers<br>• Within 2020 budget, reduced barriers to accessing tax-free childcare | • Children's protective services were moved fully to telephone or virtual services during the peak | • If meals were provided in schools or daycares, they were instructed to find a way to continue providing meals to these children |
| USA | Federal |  |  | • Reduced barriers to accessing the Special Supplemental Nutrition Program for Women, Infants and Children<br>• Coronavirus Food Assistance Program provides funding for food banks |
|  | Michigan | • Daycares were closed, except for emergency child care centres, to provide care to children of essential workers | • Transition to mixture of virtual and in-person services |  |

In Canada, daycares were closed across the country in an attempt to reduce transmission of COVID-19, a decision made by each province. As such, there were variations across provinces in how childcare for children of essential workers was provided. Most provinces either permitted some designated daycares to remain open for children of essential workers, or set up emergency childcare centres, while the British Columbia government provided financial support for essential workers to arrange in-home childcare (Government of Ontario, 2020; Government of Quebec, 2020b; Lisa Johnson, 2020; Ministry of Child and Family Development, Government of British Columbia, 2021). The Netherlands, UK and Michigan also closed daycares, but designated some to remain open for children of essential workers (Government of Michigan, 2020; “Key Worker,” 2020; Ministry of Health, Welfare and Sport, Government of the Netherlands, 2020). As daycares reopened, UK introduced policy to reduce barriers to accessing tax-free childcare (Working Families, 2020). In contrast, Australia and Singapore kept daycares open throughout the first wave, although, Australia did see many daycares close due to parents choosing to withdraw their children (Australian Bureau of Statistics, Australian Government, 2020; Channel News Asia, 2020). In response, Australia offered to pay for childcare for 2 months and also provided daycares financial support to help them maintain their staff until enrollment increased again (Australian Bureau of Statistics, Australian Government, 2020). Singapore introduced multiple policies to reduce barriers to high quality daycare, including decreasing fee caps and increasing both universal and targeted subsidies (Channel News Asia, 2020). Singapore also created the KIDStart Initiative, a pilot early childhood development program for children from low-income families (Channel News Asia, 2020).

In terms of child protective or welfare services, almost all countries transitioned to a mix of in-person and virtual services, in order to reduce in-person contact. In Ontario, these services are managed by municipal governments, creating further variation, with some municipalities continuing with fully in-person services (Government of Ontario, 2020). Only the UK had a brief period where all services were moved to fully virtual (Government of United Kingdom, 2020). Alternatively, Singapore child protective services began to proactively reach out to families in at-risk neighbourhoods, including in-person visits (Channel News Asia, 2020).

There was a large variation in the types of programs countries enacted to reduce the risk of food insecurity amongst children. Canada, the Netherlands and the USA increased funding to food banks and other emergency food relief organizations (Department of Finance Canada, 2020; U.S. Department of Agriculture, 2020; Werkgelegenheid, 2020) Several countries also instituted specific programs or interventions outside of general increased funding to food relief organizations. The USA reduced barriers to accessing existing supports, such as the Special Supplemental Nutrition Program for Women, Infants and Children (Food and Nutrition Service, U.S Department of Agriculture, 2020). Singapore created a working group to assess and address food insecurity in young families, though their recommendations had not yet been made public (Ministry of Social and Family Development, Government of Singapore, 2020). At the same time, Singapore also increased food vouchers for low-income families (Ministry of Social and Family Development, Government of Singapore, 2020). The UK mandated that any daycares and schools that were providing food to children should find a way to continue providing that food (Working Families, 2020).

Additionally, all of the countries’ governmental websites provided general information related to child mental health and well-being, though this very often focused more on adolescents. As well, all countries’ governmental websites provided links to community organizations, programs, or other supports that already exist to aid parents and young children.

## Discussion

### Overview

Our findings indicate that there was variation in policy approaches by select high-income countries in addressing maternal and early childhood concerns during the COVID-19 pandemic. Overall, Singapore introduced the greatest number of supports, along with Canada and Australia. On the other hand, Netherlands and USA introduced less. Most policies were related to financial supports. However, there was relative consensus on clinical care during the prenatal, neonatal and pediatric stages.

### Reasons for Variations in Policies

There is likely a large range of factors that influenced the variation in policies and programs introduced by the governments of these countries. The overarching political legacy and culture of these countries may have influenced the governments’ approaches. For example, within this group of countries, the USA has a legacy of less redistributive policies and cultural attitudes are less accepting of government intervention - therefore it may not be surprising that they also enacted fewer supports for parents and young children during the COVID-19 pandemic (Fishback, 2010; Stefan Svallfors, 2003). In comparison, Canada has a legacy of greater social welfare supports than the USA in the past and similarly implemented more supports for children and families during COVID-19 (Stefan Svallfors, 2003). Interestingly, the Netherlands which has a much larger social welfare support system, also introduced fewer policies to support parents and young children the COVID-19 pandemic. For example, in the Netherlands parents are already reimbursed for child care costs, depending on their income, and all employed parents have paid sick leave and additional paid “caretaking leave” to take care of a sick family member (Pfau-Effinger, 2005; Yerkes & Javornik, 2019). It may have been that the pre-pandemic support to children and families that the Netherlands provided were sufficient and additional policies were not felt to be needed.

### Potential Impacts of Policies

Preliminary studies indicated that COVID-19 and the community-wide public health measures implemented to reduce transmission of COVID-19 have already had significant impacts on maternal and child health and well-being (Public Health Ontario, 2021). COVID-19 required rapid response given the sudden disruption of long-standing systems of support, with many losing employment and access to childcare and other public health services. Furthermore, these negative impacts are likely to disproportionately affect families already at-risk, from lower socioeconomic communities (Spinelli et al., 2020). The hope is that quick implementation of the policies presented here, particularly financial supports, could mitigate some of the harms of COVID-19 public health measures. Nevertheless, the effectiveness of the policies presented here to counteract the negative impacts of COVID-19 on maternal and child health will take time to be understood.

Previous research has demonstrated the effectiveness of policies to support mothers and children in improving long-term health outcomes (Siddiqi et al., 2011). In line with the best available evidence, the OECD has compiled a list of possible governmental policies to counteract the negative impacts of COVID-19 on children (OECD, 2020b). All of the countries discussed in this study implemented some, but not all, of the recommended actions. In particular, while all governments provided information on their websites to pre-existing resources for mental health supports for children and parents, no government created new supports or policies specifically targeting this area of need.

Moreover, it should be noted that the positive impact of these policies and programs will be mediated by the extent of implementation and uptake. There is evidence that indicates that programs that automatically enroll participants and have low barriers to access will have greater uptake and impact than programs that require applications (Dorn, n.d.; Hefford et al., 2005). Thus, Singapore may see a greater impact of their financial supports due to it automatically being given to those already receiving other social supports.

### Strengths and Limitations of Study

This study used a comprehensive and iterative search strategy of the grey literature to explore this important topic. This allowed us to find policy options and programs that we had not considered a priori. It also allowed for a broad and thorough comparison of these countries’ approaches to supporting young children during the COVID-19 pandemic.

Our study is limited by the number of countries we chose to include. We purposefully chose these countries in order to provide a broad range of political traditions and COVID-19 experiences. However, since we wanted to also allow the findings to be comparable to the Canadian context, all of these countries are high-income. All of the included countries, with the exception of Singapore, are OECD comparator countries. This means that our findings may not be applicable to low- and middle-income countries. As well, though our comparison is thorough, and we briefly explored possible reasons for variations in policy choices, a much greater political analysis would be required to fully understand the reasons for each countries’ specific policy and program decisions. Lastly, while we explored each countries’ governmental response and recommendations provided by clinical colleges and bodies, the degree of uptake of these programs by the targeted population has not been explored here.

### Future Directions

In the future, the implementation and uptake of these policies and programs should be assessed. Moreover, the long-term impact of these policies and programs on health outcomes should be analyzed. Possible indicators include infant mortality rate, low birth weight rate, vaccination coverage, educational attainment and literacy rates.

## Conclusion

This study explored the variation in how families and young children are being supported during the COVID-19 pandemic in Canada, Australia, Netherlands, Singapore, UK and USA through governmental policies and programs, as well as changes to healthcare provision. All of the included countries implemented some policies and programs to support families and young children, but there was a large range in the number, size, and barriers to access these supports. None of the countries implemented supports in all of the potential areas identified. As the COVID-19 pandemic continues, there is an opportunity for every country to evaluate their overall child and family health policies and to provide additional support for families in the post-pandemic era, which will improve overall health and well-being of communities.

## Data Availability

The data that supports the findings of this study were found from public domain sources. A list of these sources can be found within the article and its supplementary materials.

