## Appendix A for "Comparative analysis of policies and programs to support families and children during COVID-19"

### **Appendix A: Search strategy**

A grey literature search was conducted for Australia, Canada, the Netherlands, Singapore, the U.K. and the U.S.A based on search strings created by Public Health Ontario librarians in custom and general web search engines. For each search, the first 100 results were reviewed and relevant articles were collected relating to COVID-19 policies to support children and mothers. Medical regulatory bodies and associations were searched without using pre-specified strings.

*Australia*

| Search Site | <https://ww.google.com.au/> with Google location changed to Australia |
| --- | --- |
|  | <https://www.google.ca/> with “Australia” added to end of the string |

Search Strings

"COVID-19" (welfare OR safety OR well-being OR wellbeing) (child OR children OR adolescent OR infant OR preschool)

"COVID-19" (welfare OR safety OR well-being OR wellbeing) (parents OR family OR household OR caregiver OR spouse impact)

"COVID-19" (prevent OR minimize OR minimise OR mitigate) (negative OR adverse OR "secondary effects" OR impact) (child OR children OR adolescent OR infant OR preschool)

"COVID-19" (prevent OR minimize OR minimise OR mitigate) (stress OR anxiety OR abuse OR neglect OR exploitation OR maltreatment) (parents OR family OR household OR caregiver OR spouse impact)

"COVID-19" ("adverse childhood experiences")(prevent OR minimize OR minimise OR mitigate)

*Canada*

| Search Sites | http://www.ophla.ca/p/customsearchcanada.html |
| --- | --- |
|  | https://cse.google.com/cse?cx=007843865286850066037:3ajwn2jlweq |
|  | <https://cse.google.com/cse/publicurl?cx=006912983573246070439:dbea-6lyv1k> |
|  | <https://www.google.ca/> with Google location changed to Canada |

Search Strings

"COVID-19" (welfare OR safety OR well-being OR wellbeing) (child OR children OR adolescent OR infant OR preschool)

"COVID-19" (welfare OR safety OR well-being OR wellbeing) (parents OR family OR household OR caregiver OR spouse impact)

"COVID-19" (prevent OR minimize OR minimise OR mitigate) (negative OR adverse OR "secondary effects" OR impact) (child OR children OR adolescent OR infant OR preschool)

"COVID-19" (prevent OR minimize OR minimise OR mitigate) (stress OR anxiety OR abuse OR neglect OR exploitation OR maltreatment) (parents OR family OR household OR caregiver OR spouse impact)

"COVID-19" ("adverse childhood experiences")(prevent OR minimize OR minimise OR mitigate)

*The Netherlands*

| Search Site | <https://ww.google.nl/> with Google location changed to the Netherlands |
| --- | --- |
|  | <https://www.google.ca/> with “Netherlands” added to end of the string |

Search Strings

"COVID-19" (welfare OR safety OR well-being OR wellbeing) (child OR children OR adolescent OR infant OR preschool)

"COVID-19" (welfare OR safety OR well-being OR wellbeing) (parents OR family OR household OR caregiver OR spouse impact)

"COVID-19" (prevent OR minimize OR minimise OR mitigate) (negative OR adverse OR "secondary effects" OR impact) (child OR children OR adolescent OR infant OR preschool)

"COVID-19" (prevent OR minimize OR minimise OR mitigate) (stress OR anxiety OR abuse OR neglect OR exploitation OR maltreatment) (parents OR family OR household OR caregiver OR spouse impact)

"COVID-19" ("adverse childhood experiences")(prevent OR minimize OR minimise OR mitigate)

Singapore

| Search Site | <https://ww.google.com.sg/> with Google location changed to Singapore |
| --- | --- |
|  | <https://www.google.ca/> with “Singapore” added to end of the string |

Search Strings

"COVID-19" (welfare OR safety OR well-being OR wellbeing) (child OR children OR adolescent OR infant OR preschool)

"COVID-19" (welfare OR safety OR well-being OR wellbeing) (parents OR family OR household OR caregiver OR spouse impact)

"COVID-19" (prevent OR minimize OR minimise OR mitigate) (negative OR adverse OR "secondary effects" OR impact) (child OR children OR adolescent OR infant OR preschool)

"COVID-19" (prevent OR minimize OR minimise OR mitigate) (stress OR anxiety OR abuse OR neglect OR exploitation OR maltreatment) (parents OR family OR household OR caregiver OR spouse impact)

"COVID-19" ("adverse childhood experiences")(prevent OR minimize OR minimise OR mitigate)

*United Kingdom*

| Search Site | <https://ww.google.co.uk/> with Google location changed to the U.K. |
| --- | --- |
|  | <https://www.google.ca/> with “(United Kingdom OR UK OR Britain)” added to end of the string |

Search Strings

"COVID-19" (welfare OR safety OR well-being OR wellbeing) (child OR children OR adolescent OR infant OR preschool)

"COVID-19" (welfare OR safety OR well-being OR wellbeing) (parents OR family OR household OR caregiver OR spouse impact)

"COVID-19" (prevent OR minimize OR minimise OR mitigate) (negative OR adverse OR "secondary effects" OR impact) (child OR children OR adolescent OR infant OR preschool)

"COVID-19" (prevent OR minimize OR minimise OR mitigate) (stress OR anxiety OR abuse OR neglect OR exploitation OR maltreatment) (parents OR family OR household OR caregiver OR spouse impact)

"COVID-19" ("adverse childhood experiences")(prevent OR minimize OR minimise OR mitigate)

*United States of America*

| Search Site | <https://ww.google.com/> with Google location changed to the U.S.A. |
| --- | --- |
|  | <https://www.google.ca/> with “(United States or US OR USA OR Michigan” added to the end of the string |

Search Strings

"COVID-19" (welfare OR safety OR well-being OR wellbeing) (child OR children OR adolescent OR infant OR preschool)

"COVID-19" (welfare OR safety OR well-being OR wellbeing) (parents OR family OR household OR caregiver OR spouse impact)

"COVID-19" (prevent OR minimize OR minimise OR mitigate) (negative OR adverse OR "secondary effects" OR impact) (child OR children OR adolescent OR infant OR preschool)

"COVID-19" (prevent OR minimize OR minimise OR mitigate) (stress OR anxiety OR abuse OR neglect OR exploitation OR maltreatment) (parents OR family OR household OR caregiver OR spouse impact)

"COVID-19" ("adverse childhood experiences")(prevent OR minimize OR minimise OR mitigate)
