## Appendix B for "Comparative analysis of policies and programs to support families and children during COVID-19"

**Appendix B: Tables with citations**

Table 1. Baseline characteristics of included countries

|  | Population (millions)*^a^* | Population Density (/km^2^)*^b^* | GDP per capita ($USD)*^a^* | % of GDP Spent on ECE*^c^* | Enrollment of children 3-5 years old in ECE (%)*^c^* | Measles immunization coverage (%)*^d^* | Gini coefficient*^e^* |
| --- | --- | --- | --- | --- | --- | --- | --- |
| Australia | 25.4 | 3 | 55 060.30 | 0.66 | 84 | 95 | 0.325 |
| Canada | 37.6 | 4 | 46 194.70 | 0.20 | 24 | 90 | 0.303 |
| Netherlands | 17.3 | 511 | 52 331.30 | 0.60 | 95 | 94 | 0.285 |
| Singapore | 5.9 | 7953 | 65 233.30 | 0.19 | 84 | 95 | 0.458 |
| UK | 66.8 | 275 | 42 330.10 | 0.65 | 100 | 91 | 0.366 |
| USA | 328.2 | 36 | 65 297.50 | 0.33 | 66.1 | 90 | 0.390 |

*^a^* as of 2019 (World Bank, 2021);

*^b^* as of 2018 (World Bank, 2021);

*^c^* for ages 0-6 years old in 2006 for Canada (OECD, 2006), for ages 4-6 years old in 2011 for Singapore (Early Childhood Development Agency, 2012), and in 2015 for all other countries (OECD, 2019);

*^d^* in 2019 (Vanderslott et al., 2013);

*^e^* in 2016 for Singapore (Li, 2020), in 2018 for all other countries (OECD, 2020)

Table 2. Comparison of epidemiology of COVID-19 in included countries until August 31, 2020

|  | Date of 1^st^ reported case*^a^* | Date of peak new cases per day*^a^* | Peak new cases per day/million*^a^* | GSI at date of peak new cases*^b^* | Testing capacity per 1000 people at peak new cases*^c^* | Test positivity at peak (%)*^c^* |
| --- | --- | --- | --- | --- | --- | --- |
| Australia | January 25 | March 30 | 15.0 | 79.17 | 2.62 | 0.7 |
| Canada | January 26 | May 4 | 47.7 | 72.69 | 0.7 | 6.6 |
| Netherlands | February 28 | April 15 | 65.4 | 79.63 | 0.33 | 21.5 |
| Singapore | January 24 | April 27 | 171.8 | 85.19 | 0.54 | 28.9 |
| UK | February 1 | April 24 | 71.4 | 79.63 | 0.37 | 19.1 |
| USA | January 21 | July 23 | 203.5 | 67.13 | 2.89 | 8.9 |

*^a^* (Ritchie et al., 2021a);

^b^ (Ritchie et al., 2021c);

*^c^* (Ritchie et al., 2021c)

Table 3. Comparison of policies regarding prenatal and pediatric care in selected countries during COVID-19

| Country | Jurisdiction | Prenatal Care | Well-baby visit schedule | Vaccines |
| --- | --- | --- | --- | --- |
| Australia | Federal | - Continue routine antenatal care, though provider can arrange for extra scans if COVID-19 positive^a^ | - Continue routine schedule with mix of virtual and in-person visits^b^ | - Recommendation to continue vaccinations as scheduled^c^ |
| Canada | Federal | - Modified schedule, with mix of virtual and in-person practice^d^ - Consider delaying routine appointments for pregnant patients being tested or COVID-19 positive^e^ | - Modified schedule with mix of virtual and in-person visits^d^ | - Recommendation to continue vaccinations as scheduled^f^ |
|  | Alberta | - Prenatal classes suspended^g^ | - Public health nurse or midwife to continue postpartum care as usual^g^ - Physician visits should continue, but may be virtual depending on location^g^ | - Routine immunizations to continue, with exception of school program^h^ |
|  | British Columbia | - Reduced antenatal visits, with a mix of virtual and in-person practice^i^ | - At the discretion of physician^j^ | - Routine immunization schedule to continue^k^ |
|  | Ontario | - Modified prenatal visit schedule^d^ | - Modified well-child schedule, with mix of in-person and virtual^d^ | - Routine infant vaccination schedule^d^ - Consider delaying 4-6 year old immunizations^d^ |
|  | Quebec | - Modified schedule, with mix of virtual and in-person practice^rl^ | - Routine schedule to be continued, though may be virtual^l^ | - Routine infant vaccination schedule^m^ |
| Netherlands | | - Continue routine schedule^n^ | - Combination of in person and virtual visits^o^ - In-person weight checks by appointment only if there are concerns^o^ | - Routine schedule to continue^o^ |
| Singapore | | - Postpone non-critical appointments if on a Stay-Home Notice or quarantine^p^ - Otherwise, continue routine schedule^p^ |  | - Continue routine schedule^q^ |
| UK | | - Continue routine antenatal care, although can be modified, unless suspected or confirmed COVID-19^r^ | - Continue routine 6-8 week infant examination^s^ | - Continue routine childhood vaccinations as scheduled^s^ |
| USA | Federal | - Continue to provide medically necessary prenatal care, referrals and consultations but can modify/reduce if risk outweighs benefit^t^ | - Continue with routine well-baby visits^u^ | - Recommendation to continue infant and toddler vaccinations as scheduled^u^ |
|  | Michigan | - Policy varied by health service provider^v^ - General reduction in in-person visits^v^ | - At the discretion of physician^w^ | - Continue routine schedule^x^ |

^a^(The Royal Australian and New Zealand College of Obstetricians and Gynaecologists, 2020); ^b^(NSW Health, 2020b); ^c^(NSW Health, 2020a); ^d^(Bogler & Bogler, 2020); ^e^(Audibert et al., 2020); ^f^(Canadian Paediatric Society, 2020); ^g^(Alberta Health Services, 2020a);^h^(Alberta Health Services, 2020b); ^i^(Provincial Health Services Authority, 2020); ^j^(Health Link BC, 2020b); ^k^(Health Link BC, 2020a); ^l^(Government of Quebec, 2020a); ^m^(Government of Quebec, 2020b); ^n^(Nederlandese Vereniging Voor Obstetrie & Gynaecologie, 2020); ^o^(Jong JGZ, 2020); ^p^(College of Obstetricians & Gynaecologists, Singapore, 2020); ^q^(Health Promotion Board, Government of Singapore, 2020); ^r^(Royal College of Obstetricians & Gynaecologists, 2020); ^s^(Santhanam, 2020); ^t^(American College of Obstetricians and Gynecologists, 2020); ^u^(American Academy of Pediatrics, 2021); ^v^(Government of Michigan, 2020a); ^w^(Government of Michigan, 2020b); ^x^(Michigan Department of Health and Human Services, 2020)

Table 4. Comparison of additional maternal supports offered by governments in response to COVID-19 in selected countries

| Country | Jurisdiction | Financial Supports | Domestic Violence and Housing | Other |
| --- | --- | --- | --- | --- |
| Australia | Federal | - One-time payment of $750(AUD) to anyone who receives Family Tax Benefit^a^ | - $150 million (AUD) to support community organizations addressing domestic violence^b^ |  |
| Canada | Federal | - One-time top-up of $300 (CAD) for Canada Child Benefit per child^c^ - Creation of Canada Recovery Caregiving Benefit, to provide income support for parents that must stay home to care for sick children during COVID-19^d^ | - Creation of new shelters for Indigenous women and children^e^ - Increased financial support of women’s shelters^e^ - Virtual domestic violence supports for military personnel^f^ | - Funded research on the social impacts of COVID-19 on children and families^g^ |
|  | Alberta |  |  |  |
|  | British Columbia | - Additional $225/month (CAD) for children with special needs^h^ |  |  |
|  | Ontario | - One-time payment of $200-$250(CAD) per child^i^ | - Increased funding to support victims of domestic violence^j^ |  |
|  | Quebec |  |  |  |
| Netherlands | |  | - Country wide media campaign with information about domestic violence^k^ | - Funding research on the impact of COVID-19 on maternal mental health^l^ |
| Singapore | | - One-time payment of $1000(SGD) to low-income families affected by COVID-19^m^ - Increased child benefit by $300(SGD) for each parent in household for one month^n^ - One-time additional support for newborns, in order to encourage families to have children during COVID-19^o^ |  |  |
| UK | |  | - Introduced laws strengthening protections and increasing assistance to those experiencing domestic violence (was already underway, completed during COVID-19)^p^ | - Funding research on the impact of COVID-19 on maternal mental health^q^ |
| USA | Federal | - No change to federal Child Tax Credit^r^ | - CARES Act includes $% million for emergency shelter via the Family Violence Prevention and Services Act^s^ |  |
|  | Michigan |  |  |  |

^a^(Department of Social Services, Australian Government, 2020); ^b^(Murphy, 2020); ^c^(Canada Revenue Agency, Government of Canada, 2020a); ^d^(Canada Revenue Agency, Government of Canada, 2020b); ^e^(Department of Finance Canada, Government of Canada, 2020); ^f^(Canadian Armed Forces, 2020b); ^g^(Canadian Institutes of Health Research, Government of Canada, 2020); ^h^(Ministry of Children and Family Development, Government of British Columbia, 2020); ^i^(Ministry of Education, Government of Ontario, 2020); ^j^(Government of Ontario, 2020a); ^k^(Government of Netherlands, 2020); ^l^(Dutch Research Council (NWO), 2020); ^m^(Ministry of Social and Family Development, Government of Singapore, 2020b); ^n^(Medina, 2020); ^o^(Budget 2020, Government of Singapore, 2020b); ^p^(Home Office, UK Government, 2020); ^q^(Staniscuaski et al., 2020); ^r^(Marr et al., 2020); ^s^(National Network to End Domestive Violence, 2020)

Table 5. Comparison of additional supports for childcare and early childhood development by governments in response to COVID-19 in selected countries

| Country | Jurisdiction | Daycares and Childcare | Child Protective Services | Food Security |
| --- | --- | --- | --- | --- |
| Australia | Federal | - Offered free childcare from April to July, 2020 during COVID-19^a^ - No official closure of daycares, though many closed as parents withdrew children^b^ - Financial support for childcare centres^b,c^ | - Transition to mixture of virtual and in-person services^d^ | - Increased funding for emergency food relief organizations^e^ |
| Canada | Federal | - Emergency family care during COVID for military families^f^ |  | - $100 million in funding for food banks and local food organizations^g^ |
|  | Alberta | - Daycares were closed, except for emergency child care centres for children of essential workers^h^ | - Child intervention services not open to public, only available by phone^i^ |  |
|  | British Columbia | - Daycares were closed^j^ - Temporary Emergency Relief funding was provided to daycare centres to allow for them to retain staff and maintain spots for children when they reopen^j^ - Extra supports for children of essential workers, to allow for in-own-home childcare^j^ - Affordable Child Care Benefit continued, even if child was not able to attend daycare^j^ | - Transition to mixture of virtual and in-person services^j^ | - Various grants available^k^ |
|  | Ontario | - Daycares were closed, except for emergency child care centres, to provide care to children of essential workers^l^ | - Transition to mixture of virtual and in-person services^m^ |  |
|  | Quebec | - Daycares were closed, except for emergency child care centres, to provide care to children of essential workers^n^ |  |  |
| Netherlands | | - Daycares closed, except for children of essential workers^o^ - Continued payments for child-care, even if child care centres were closed^o^ | - Mixture of virtual and in-person services^p^ | - Increased funding for food banks^q^ |
| Singapore | | - Increased already existing universal and targeted subsidies for childcare^r^ - Lowered fee caps on childcare, in order to make high-quality childcare more affordable^r^ - Increased supports for children in pre-school with special needs^r^ - Started KIDStart Initiative, a pilot project for children from low-income families^r^ | - Children’s protective services proactively reaching out to at-risk families, including continued in-person visits^r^ | - Created a working group to assess and address food insecurity in young families during COVID-19^s^ - Increased food vouchers for low-income families^t^ |
| UK | | - Daycares were closed, except for those of essential workers^u^ - Within 2020 budget, reduced barriers to accessing tax-free childcare^v^ | - Children’s protective services were moved fully to telephone or virtual services during the peak^w^ | - If meals were provided in schools or daycares, they were instructed to find a way to continue providing meals to these children^x^ |
| USA | Federal |  |  | - Reduced barriers to accessing the Special Supplemental Nutrition Program for Women, Infants and Children^y^ - Coronavirus Food Assistance Program provides funding for food banks^y^ |
|  | Michigan | - Daycares were closed, except for emergency child care centres, to provide care to children of essential workers^z^ | - Transition to mixture of virtual and in-person services^aa^ |  |

^a^(Australian Bureau of Statistics, Australian Governme`nt, 2020); ^b^(Prime Minister of Australia, 2020); ^c^(Department of Education, Skills and Employment, Government of Australia Centre, 2020); ^d^(Department for Child Protection, 2020); ^e^(Murphy, 2020); ^f^(Canadian Armed Forces, 2020a); ^g^(Department of Finance Canada, Government of Canada, 2020); ^h^(Lisa Johnson, 2020); ^i^(Government of Alberta, 2020); ^j^(Ministry of Child and Family Development, Government of British Columbia, 2021); ^k^(BC Food Security Gateway, 2020); ^l^ (Ministry of Health, Government of Ontario, 2020); ^m^(Government of Ontario, 2020b); ^n^(Government of Quebec, 2020c); ^o^(Ministry of Health, Welfare and Sport, Government of the Netherlands, 2020); ^p^(Jeugdzord Nederland, 2020); ^q^(Werkgelegenheid, 2020); ^r^(Channel News Asia, 2020); ^s^(Ministry of Social and Family Development, Government of Singapore, 2020a); ^t^(Budget 2020, Government of Singapore, 2020a); ^u^(“Key Worker,” 2020); ^v^(HM Treasury, UK Government, 2020); ^w^(Government of United Kingdom, 2020); ^x^(Working Families, 2020); ^y^(Food and Nutrition Service, U.S Department of Agriculture, 2020); ^z^(The Office of Governor Gretchen Whitmer, Government of Michigan, 2020); ^aa^(Burgio, 2020)

Ontario Public Health Association. (2020, October 30). *Your COVID-18 Summary for Oct. 30th—National Projections* [Personal communication].

Prime Minister of Australia. (2020, April 2). *Early childhood education and care relief package*. https://www.pm.gov.au/media/early-childhood-education-and-care-relief-package

Provincial Health Services Authority. (2020). *Antenatal visits during COVID-19 pandemic*. http://www.bccdc.ca/Health-Professionals-Site/Documents/COVID19_AntenatalVisitsDuringPandemic.pdf.

Ritchie, H., Ortiz-Ospina, E., Beltekian, D., Mathieu, E., Hasell, J., Macdonald, B., Giattino, C., & Roser, M. (2021a). *Coronavirus (COVID-19) Cases—Statistics and Research*. Our World in Data. https://ourworldindata.org/covid-cases

Ritchie, H., Ortiz-Ospina, E., Beltekian, D., Mathieu, E., Hasell, J., Macdonald, B., Giattino, C., & Roser, M. (2021b). *Coronavirus (COVID-19) Testing—Statistics and Research*. Our World in Data. https://ourworldindata.org/coronavirus-testing

Ritchie, H., Ortiz-Ospina, E., Beltekian, D., Mathieu, E., Hasell, J., Macdonald, B., Giattino, C., & Roser, M. (2021c). *Policy Responses to the Coronavirus Pandemic—Statistics and Research*. Our World in Data. https://ourworldindata.org/policy-responses-covid

Royal College of Obstetricians & Gynaecologists. (2020). *Coronavirus (COVID-19) Infection in Pregnancy—Version 12*. https://www.rcog.org.uk/globalassets/documents/guidelines/2020-10-14-coronavirus-covid-19-infection-in-pregnancy-v12.pdf

Santhanam, L. (2020). *Postnatal Maternal and Infant Care during the COVID-19 Pandemic: A guide for General Practice (Version 3)*. Royal College of General Practitioners. https://elearning.rcgp.org.uk/pluginfile.php/148864/mod_page/content/86/Postnatal%20Maternal%20and%20Infant%20Care%20during%20the%20COVID-19%20Pandemic%20-%20A%20guide%20for%20General%20Practice%20Version%203%20%2811.11.2020%29.pdf

Staniscuaski, F., Reichert, F., Werneck, F. P., de Oliveira, L., Mello-Carpes, P. B., Soletti, R. C., Almeida, C. I., Zandona, E., Ricachenevsky, F. K., Neumann, A., Schwartz, I. V. D., Tamajusuku, A. S. K., Seixas, A., Kmetzsch, L., & Parent in Science Movement†. (2020). Impact of COVID-19 on academic mothers. *Science*, *368*(6492), 724.1-724. https://doi.org/10.1126/science.abc2740

The Office of Governor Gretchen Whitmer, Government of Michigan. (2020, April 15). *Whitmer—Executive Order 2020-51: Expanding child care access during the COVID-19 pandemic—RESCINDED*. https://www.michigan.gov/whitmer/0,9309,7-387-90499_90705-526011--,00.html

The Royal Australian and New Zealand College of Obstetricians and Gynaecologists. (2020, August 6). *Info for Pregnant Women*. https://ranzcog.edu.au/statements-guidelines/covid-19-statement/information-for-pregnant-women

Vanderslott, S., Dadonaite, B., & Roser, M. (2013, May 10). *Vaccination*. Our World in Data. https://ourworldindata.org/vaccination

Werkgelegenheid, M. van S. Z. en. (2020, March 24). *Noodsteun om voedselbanken draaiende te houden—Nieuwsbericht—Rijksoverheid.nl* [Nieuwsbericht]. Ministerie van Algemene Zaken. https://www.rijksoverheid.nl/actueel/nieuws/2020/03/24/noodsteun-om-voedselbanken-draaiende-te-houden

Working Families. (2020). *Coronavirus (COVID-19) – What financial support is there for working families?* Working Families. https://workingfamilies.org.uk/articles/coronavirus-support/

World Bank. (2021). *World Bank Open Data | Data*. Open Data. https://data.worldbank.org/
